# “The Effect of Inter-City Travel Restrictions on Geographical Spread of COVID-19: Evidence from Wuhan, China”

**DOI:** 10.1101/2020.04.16.20067504

**Authors:** Billy J. Quilty, Charlie Diamond, Yang Liu, Hamish Gibbs, Timothy W. Russell, Christopher I. Jarvis, Kiesha Prem, Carl A.B. Pearson, Samuel Clifford, Stefan Flasche, CMMID COVID-19 working group, Petra Klepac, Rosalind M. Eggo, Mark Jit

## Abstract

**Background:** To contain the spread of COVID-19, a *cordon sanitaire* was put in place in Wuhan prior to the Lunar New Year, on 23 January 2020, restricting travel to other parts of China. We assess the efficacy of the *cordon sanitaire* to delay the introduction and onset of local transmission of COVID-19 in other major cities in mainland China.

**Methods:** We estimated the number of infected travellers from Wuhan to other major cities in mainland China from November 2019 to March 2020 using previously estimated COVID-19 prevalence in Wuhan and publicly available mobility data. We focused on Beijing, Chongqing, Hangzhou, and Shenzhen as four representative major cities to identify the potential independent contribution of the *cordon sanitaire* and holiday travel. To do this, we simulated outbreaks generated by infected arrivals in these destination cities using stochastic branching processes. We also modelled the effect of the *cordon sanitaire* in combination with reduced transmissibility scenarios representing the effect of local non-pharmaceutical interventions.

**Findings:** In the four cities, given the potentially high prevalence of COVID-19 in Wuhan between Dec 2019 and early Jan 2020, local transmission may have been seeded as early as 2 - 8 January 2020. By the time the *cordon sanitaire* was imposed, simulated case counts were likely in the hundreds. The *cordon sanitaire* alone did not substantially affect the epidemic progression in these cities, although it may have had some effect in smaller cities.

**Interpretation:** Our results indicate that the *cordon sanitaire* may not have prevented COVID-19 spread in major Chinese cities; local non-pharmaceutical interventions were likely more important for this.

**Research in Context:** *Evidence before this study:* In late 2019, severe acute respiratory syndrome coronavirus 2 (SARS-CoV-2) was detected in Wuhan, China. In response to the outbreak, authorities enacted a *cordon sanitaire* in order to limit spread. Several studies have sought to determine the efficacy of the policy; a search of PubMed for “coronavirus AND (travel restrictions OR travel ban OR shutdown OR cordon sanitaire) AND (Wuhan OR China)” returned 24 results. However other studies have relied on reported cases to determine efficacy, which are likely subject to reporting and testing biases. Early outbreak dynamics are also subject to a significant degree of stochastic uncertainty due to small numbers of cases.

*Added value of this study:* Here we use publicly-available mobility data and a stochastic branching process model to evaluate the efficacy of the *cordon sanitaire* to limiting the spread of COVID-19 from Wuhan to other cities in mainland China, while accounting for underreporting and uncertainty. We find that although travel restrictions led to a significant decrease in the number of individuals leaving Wuhan during the busy post-Lunar New Year holiday travel period, local transmission was likely already established in major cities. Thus, the travel restrictions likely did not affect the epidemic trajectory substantially in these cities.

*Implications of all the available evidence:* A *cordon sanitaire* around the epicentre alone may not be able to reduce COVID-19 incidence when implemented after local transmission has occurred in highly connected neighbors. Local non-pharmaceutical interventions to reduce transmissibility (e.g., school and workplace closures) may have contributed more to the observed decrease in incidence in mainland China.

## Introduction

Since late 2019, severe acute respiratory syndrome coronavirus 2 (SARS-CoV-2), the causative agent of Coronavirus Disease 2019 (COVID-19), has spread to over 114 countries and was declared a pandemic on 11 March 2020(1). Some countries have enacted *cordon sanitaire*-type travel restrictions, either to prevent the export of infections from an initial disease epicentre (such as Wuhan in January 2020 (2) or Northern Italy in March 2020 (3)) to other countries and regions, or to prevent the import of infections from high-risk countries or regions (such as the USA’s ban on travel from Europe (4)). *Cordons sanitaires* aim to curb the number of infected travellers entering a region with a high proportion of susceptible individuals, where they may seed additional chains of transmission. However, they are often reactive, occurring only after a significant number of transmission chains have been established. Hence, the efficacy of *cordons sanitaires* in averting or delaying outbreaks in other locations is an open question.

Chinese authorities imposed a *cordon sanitaire* on the city of Wuhan on 23 Jan 2020 (2) and extended the travel restrictions to the whole of Hubei province by 26 Jan 2020 (5). The restrictions were imposed one day prior to the Lunar New Year holidays and during *Chunyun*, the 40-day holiday travel period that marks the largest annual human migration event in the world (6). At the same time, other public health interventions, such as physical distancing, were also enacted across China (7).

This study aims to assess the impacts of the *cordon sanitaire* around Wuhan, the epicentre of the COVID-19 pandemic, on reducing incidence and delaying outbreaks in other well-connected large population centres in mainland China. We used publicly available mobility data based on Location Based Service (LBS) provided by Baidu Huiyan, to construct four mobility scenarios. Combined with daily estimated prevalence of COVID-19 in Wuhan before 11 February 2020 by Kucharski et al. (8), we simulated the daily importations of infected travellers to Beijing, Chongqing, Hangzhou and Shenzhen to assess the risk that they would cause sustained local transmission.

## Methods

### Estimating number of infected travellers

We obtained daily prefecture-level human mobility data, expressed by a relative index scale, for mainland China from Baidu Huiyan for both the 2019 and 2020 travel periods surrounding the LNY, known as *Chunyun*. The platform aggregates mobile phone travel data from an estimated 189 million daily active users, processing >120 billion daily positioning requests mainly through WiFi and GPS (9).

We examined the proportions of the total outflow leaving Wuhan and entering all other prefectures in China (excluding Wuhan). We then selected Beijing, Chongqing, Hangzhou and Shenzhen for further analysis as major population centres with substantial travel with Wuhan and a wide geographic spread. We assume that the early transmission dynamics of SARS-CoV-2 in cities of this size were similar to that in Wuhan.

To estimate the absolute number of daily travellers leaving Wuhan we assumed that each unit of Baidu’s migration index corresponds linearly to 50,000 travellers. This was chosen as the most credible value after synthesising evidence from several sources (7,10–13) (see Supplementary Appendix 1).

We calculated the total number of daily travellers leaving Wuhan and entering each city by taking the product of the scaling factor, the total daily outflow index from Wuhan, and the daily proportion of travellers from Wuhan entering the four cities. From the daily number of arrivals, we then simulated the daily number of infected travellers by drawing from a Poisson process based on COVID-19 prevalence among Wuhan residents (Supplementary Appendix 2). Daily estimated COVID-19 prevalence in Wuhan was retrieved from outputs of a published model on the early dynamics of COVID-19 transmission in Wuhan (8). We assumed that individuals would travel regardless of their infection status, and Wuhan was the sole source of infected individuals.

We examined four travel scenarios (Table 1), Scenario 1 is based on the observed travel pattern in 2020, and represents the *Chunyun* period with *cordon sanitaire* introduced on 23 January. Scenario 2 represents a counterfactual travel pattern used to evaluate how the COVID-19 outbreak would spread if no *cordon sanitaire* was implemented. This was based upon the actual travel from Wuhan for the equivalent *Chunyun* period in 2019. In Scenario 3, we synthesized a hypothetical travel pattern to represent a typical non-*Chunyun* period with *cordon sanitaire* introduced on 23 January, using outward travel flow on representative non-*Chunyun* days in 2019. Scenario 4 is a variation on Scenario 3 in which no *cordon sanitaire* was implemented.

**Table 1.** Scenarios describing different possible travel patterns out of Wuhan used in simulations.

| Scenario | Time of year | Source year | *Cordon sanitaire* imposed | Observed/hypothetical |
| --- | --- | --- | --- | --- |
| 1 | *Chunyun* | 2020 | Yes | Observed |
| 2 | *Chunyun* | 2019 | No | Observed |
| 3 | Non-*Chunyun* | 2019 & 2020 | Yes | Hypothetical |
| 4 | Non-*Chunyun* | 2019 | No | Hypothetical |

We extended the corresponding outflow time series to to the early stages of the outbreak (22 November 2019), by assuming the outflow from Wuhan to equal to the average daily outflow on representative non-*Chunyun* days, whilst accounting for weekday effects. The pairwise travel flow proportions between Wuhan and each other prefecture-level city was only available between 1 January - 1 March, 2020, so an approximation of the general flow magnitude was used for dates outside of the observed range (22 November - 31 December) and in simulated aspects of our scenarios i.e. *Chunyun* affected travel days in non-*Chunyun* scenarios. A more detailed description of how each scenario was formulated is in Supplementary Appendix 3.

### Branching process transmission model

As cases in China during the early epidemic were likely underreported (14), we used a stochastic branching process model to simulate outbreaks in each of the four cities. Consistent with the prevalence estimates from Wuhan (8), we began simulating travel from Wuhan on 22 November 2019, and calculated incidence up to 1 March 2020. For each simulated infected arrival in each city on a given day, an independent branching process is generated, with:

- A negative binomial offspring distribution with a time-varying mean effective reproduction number (R_e_) with baseline 2.2 (15) and overdispersion (*k*, variability in the number of secondary cases resulting from an infected case) of 0.54 (15).
- A discretized log-normal serial interval with mean of 4.7 and standard deviation of 2.9 (16).

We assume that in the initial phases of the epidemic (prior to the cordon sanitaire), the effective daily reproduction number (R_e_) was 2.2 (15,17). The date at which the probability of sustained transmission exceeded a threshold of 95% (i.e, an outbreak occurring) given R_e_ of 2.2 and k = 0.54 was used to evaluate the effect of travel restrictions (details in Supplementary Appendix 4). A sensitivity analysis for k using SARS-like (0.16) and H1N1-like (2.0) overdispersion in R_e_ is shown in Table S3. To simulate the effect of local non-pharmaceutical intervention measures (NPIs) such as physical distancing and workplace and school closures in addition to travel restrictions (18), we compare R_e_=2.2 in the absence of interventions (no change, unmitigated local outbreak), to 1.1 (50% reduction, slowing epidemic, R_e_>1), or 0.55 (75% reduction, suppressing epidemic, R_e_<1). We assume additional interventions took effect on the same date as the introduction of the *cordon sanitaire*, 23 January 2020.

### Implementation

All analyses were carried out using R version 3.6.2. The branching process model was implemented using the package *projections* version 0.4.1 (19).

## Results

### Effect of the *cordon sanitaire* on mobility

A gradual increase in the outflow from Wuhan in the weeks prior to the LNY was observed in both 2020 and 2019, exemplifying the *Chunyun* period (Figure 1). Comparing the 23 days prior to the introduction of the *cordon sanitaire* in scenarios 1 and 2, we estimate daily outflow was 21.7% (95% CI 9.78% - 33.6%) higher in 2020 than the equivalent period in 2019. A surge in volume in the 3 days preceding the *cordon sanitaire* can be seen in scenario 1 (2020), where an estimated 1.69 million left Wuhan, in line with other estimates (7). A similar outflow immediately before the LNY observed in scenario 2 (2019) suggests the surge cannot necessarily be attributed to upcoming travel restrictions. This is further reflected by the 22.5% between-year increase during this 3-day window not being substantially greater than the average daily outflow increase.

**Figure 1.**
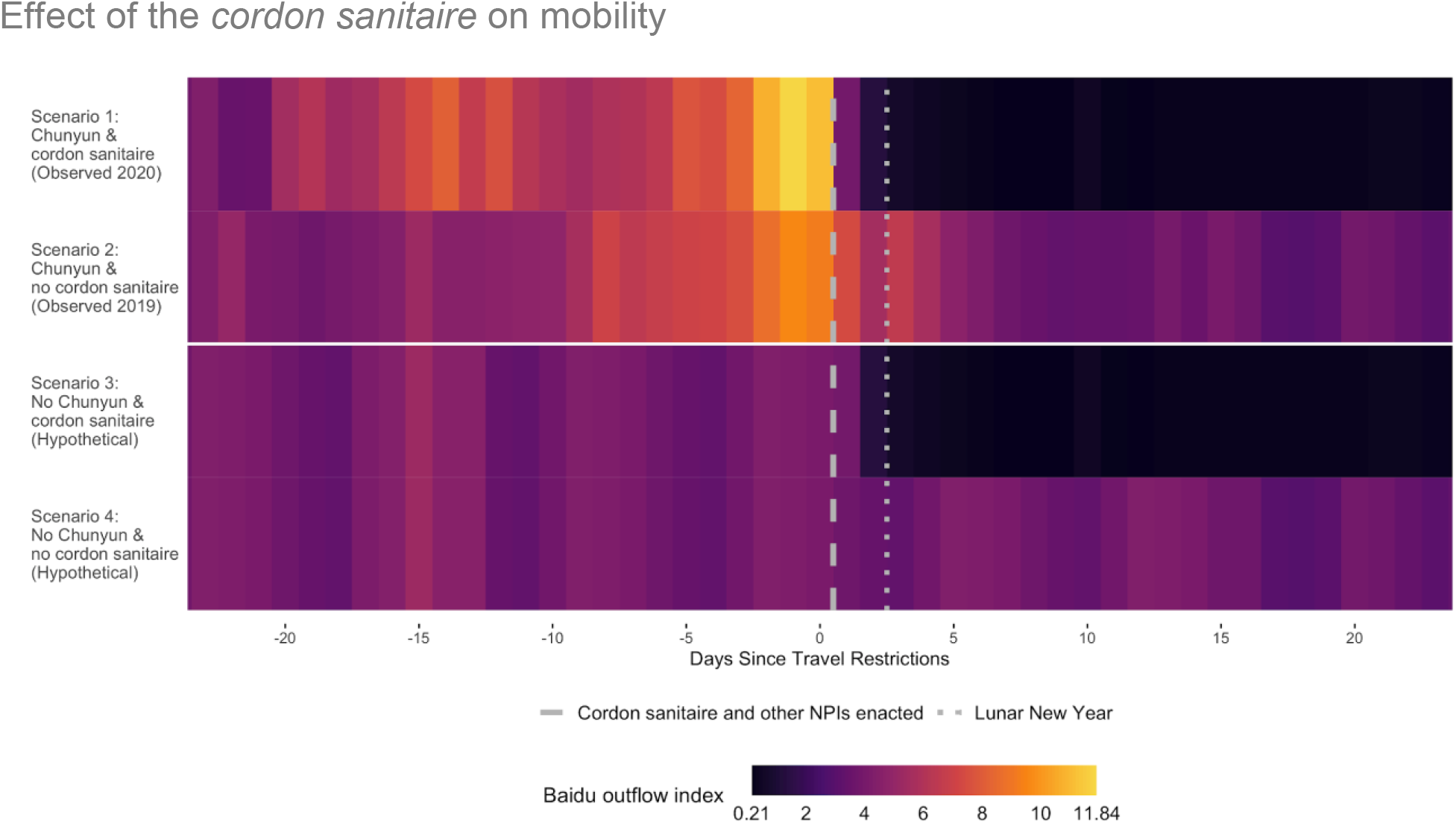
Total domestic travel outflow from Wuhan under 4 travel pattern scenarios. Dates presented on a scale in relation to the introduction of the cordon sanitaire (0 = January 23rd).

The *cordon sanitaire* had a stark effect on reducing the total outflow from Wuhan. Comparing the mean daily outflow in the 23 days preceding restrictions with the 23 days after, volume fell by 92.7%, from 345,000 (95% CI 299,000 - 390,000) average daily travellers to 25,300 (95% CI 8,590 - 42,000). In comparison, volume fell by 30.2% during the equivalent period in 2019 from 290,000 (95% CI 252,000 - 328,000) to 203,000 (95% CI 177,000 - 228,000). After restrictions were imposed, travel volume declined to a low plateau over 5 days, during which approximately 330,000 people left. On the lowest day (3 February) we estimate 10,500 people left Wuhan, which likely represents only essential journeys.

In our hypothetical scenarios, we simulated the outbound flow with the additional travel volume due to *Chunyun* removed. By comparing scenarios 2 and 4 during *Chunyun* (10 January - 18 February, 2020) we estimate that 60,000 (95% CI 32,000 - 88,100) extra travellers left Wuhan every day because of *Chunyun*.

We found that in all but one prefecture with over 7 million inhabitants, the *cordon sanitaire* on 23 January did not substantially change the time at which sustained transmission was likely to occur (Figure S1), but the picture was more mixed in smaller cities. Of the four representative major cities selected for further analysis, during their pre-restriction travel phase in Scenario 1 (1 January - 23 January, 2020): Beijing experienced a high volume of travel with approximately 1,510 (95% CI 1,200 - 1,820) mean daily travellers from Wuhan; Chongqing had the highest at 1650 (95% CI 1,320 - 1,970); Hangzhou received relatively fewer with 451 (95% CI 362 - 541); and Shenzhen had a medium travel volume from Wuhan with 820 (95% CI 664 - 976) mean daily travellers (Figure 2A).

**Figure 2.**
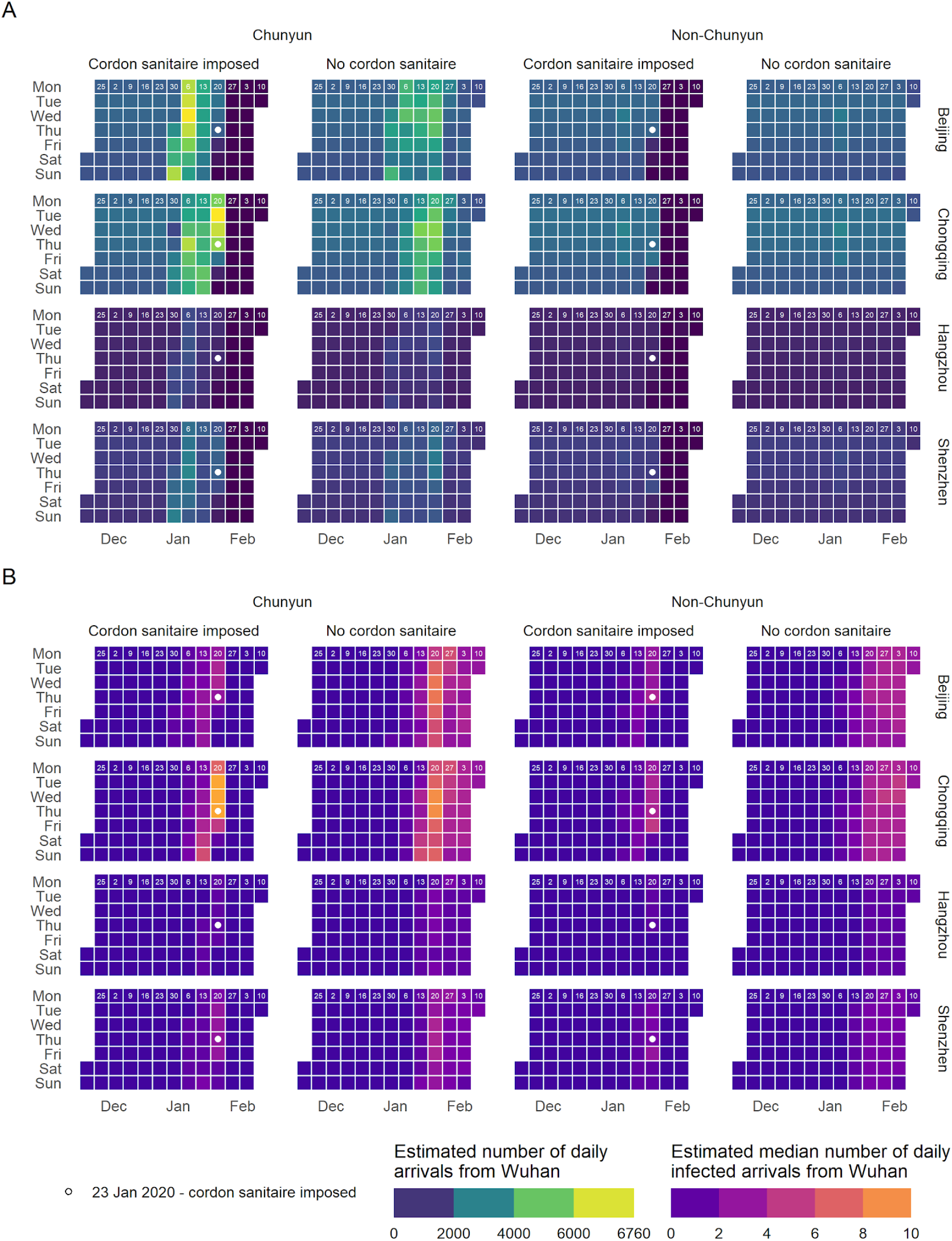
A. Estimated number of daily arrivals from Wuhan for the four chosen cities (Beijing, Chongqing, Hangzhou and Shenzhen, top to bottom) and four scenarios (1-4, left to right). Numerical day of the month given for each Monday of the week in each timeline in white. Cordon sanitaire was imposed on 23 Jan (white dot). B. Estimated median number of daily **infected** arrivals from Wuhan for the four chosen cities (Beijing, Chongqing, Hangzhou and Shenzhen, top to bottom) and four scenarios (1-4, left to right). Numerical day of the month given for each Monday of the week in each timeline in white. Cordon sanitaire was imposed inWuhan on 23 Jan (white dot).

### Effect of the *cordon sanitaire* on importations of infected persons to other major Chinese cities

We estimate that infected individuals began arriving on a daily basis in other major population centres in early January in Scenario 1 (observed *Chunyun* travel profile and *cordon sanitaire imposed*) (Figure 2B). The estimated number of infected arrivals on a given day peaked on the day before the travel restrictions at 4 (95% uncertainty interval (UI) 1 - 8) in Beijing, 10 (95% UI 5 - 17) in Chongqing, 1 (95% UI 0 - 4) in Hangzhou and 3 (95% UI 1 - 7) in Shenzhen. Travel restrictions reduced the number of infected arrivals to below 1 in all four cities within two days (Figure 2B). In Scenario 2 (*Chunyun* travel profile without *cordon sanitaire*), the number of daily infected arrivals decreases slightly after the *Chunyun* travel period, then stabilizes (Figure 2B). In cities with populations below 7 million, infected individuals began arriving later, so the *cordon sanitaire* may have acted to delay or prevent the arrival of infected individuals (Figure S1).

In Scenario 3 (*Non-Chunyun* with travel restrictions) the estimated number of daily infected arrivals is marginally lower than Scenario 1, peaking at 3 (95% UI 1 - 8) in Beijing, 4 (95% UI 5 - 9) in Chongqing, 1 (95% UI 0 - 4) in Hangzhou and 3 (95% UI 0 - 7) in Shenzhen.

### Effect of the *cordon sanitaire* on outbreaks in other major Chinese cities

Due to the volume of outbound travel from Wuhan in Scenario 1, we estimate that sustained local transmission was likely to have already occurred in the four cities in early January, several weeks prior to the introduction of the *cordon sanitaire* (Table 2). On the date travel restrictions from Wuhan were imposed, local infections were likely to already have reached over 500 in Beijing and Chongqing (Table 3). Outbreaks started later and were smaller on the date of the shutdown in Hangzhou and Shenzhen compared to Beijing and Chongqing, which reflects the relative volume of travel from Wuhan (Figure 2A).

**Table 2.**
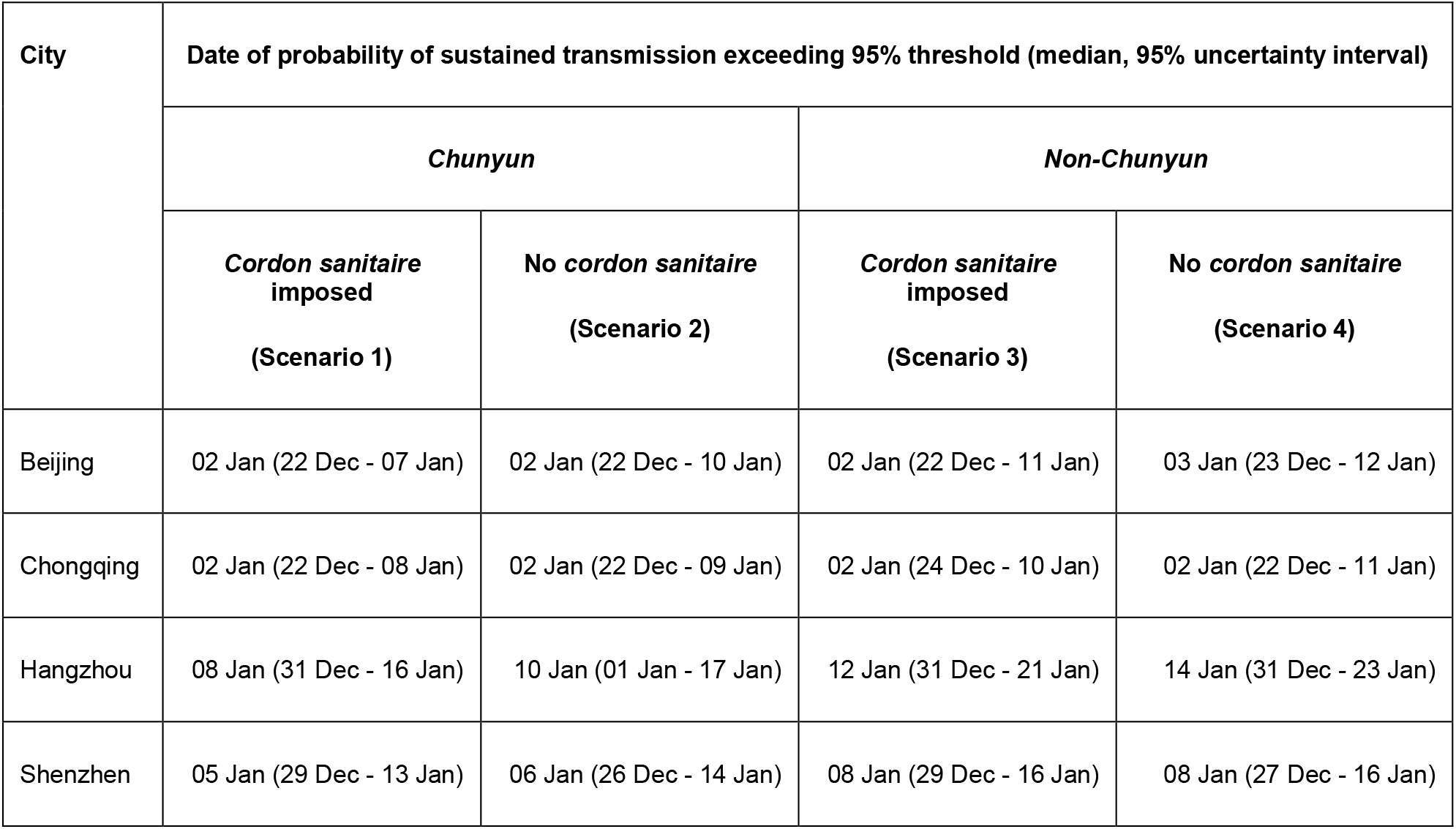
Dates at which the probability of sustained transmission breached the 95% threshold for the four prefecture-level cities of interest in each of the 4 scenarios.

| City | Date of probability of sustained transmission exceeding 95% threshold (median, 95% uncertainty interval) |  |  |  |
| --- | --- | --- | --- | --- |
|  | Chunyun |  | Non-Chunyun |  |
|  | Cordon sanitaire imposed<br>(Scenario 1) | No cordon sanitaire<br>(Scenario 2) | Cordon sanitaire imposed<br>(Scenario 3) | No cordon sanitaire<br>(Scenario 4) |
| Beijing | 02 Jan (22 Dec - 07 Jan) | 02 Jan (22 Dec - 10 Jan) | 02 Jan (22 Dec - 11 Jan) | 03 Jan (23 Dec - 12 Jan) |
| Chongqing | 02 Jan (22 Dec - 08 Jan) | 02 Jan (22 Dec - 09 Jan) | 02 Jan (24 Dec - 10 Jan) | 02 Jan (22 Dec - 11 Jan) |
| Hangzhou | 08 Jan (31 Dec - 16 Jan) | 10 Jan (01 Jan - 17 Jan) | 12 Jan (31 Dec - 21 Jan) | 14 Jan (31 Dec - 23 Jan) |
| Shenzhen | 05 Jan (29 Dec - 13 Jan) | 06 Jan (26 Dec - 14 Jan) | 08 Jan (29 Dec - 16 Jan) | 08 Jan (27 Dec - 16 Jan) |

**Table 3.** Estimated number of local infections in each of the four cities of interest on 23 January 2020, the date the cordon sanitaire was imposed.

| Prefecture-level city | Number of locally transmitted infections by 23 Jan (cumulative) (median, 95% uncertainty interval) |
| --- | --- |
| Beijing | 615 (143 - 9120) |
| Chongqing | 560 (112 - 9400) |
| Hangzhou | 112 (2 - 3550) |
| Shenzhen | 281 (10 - 6800) |

No substantial difference was observed in the daily incidence in the scenarios with and without travel restrictions in the four cities after the *cordon sanitaire* was imposed on 23 January. By then there were enough infected people to sustain local transmission without imported infections (Figure 3). In an unmitigated outbreak where R_e_ remains at 2.2, incidence continues to increase exponentially. Reducing the effective reproduction number *R*_e_ to simulate local control measures to reduce transmission did have a substantial effect; by 1 March, in Scenario 1 (*Chunyun* and *cordon sanitaire*) when *R*_e_ was reduced by 75% to 0.55, the median daily incidence was 1 (95% CI 0 - 42) infections in Beijing, 2 (95% CI 0 - 31) infections in Chongqing, 0 (95% CI 0 - 9) infections in Hangzhou, and 0 (95% CI 0 - 11) infections in Shenzhen, which is comparable to the actual observed case counts on 1 March from the WHO Situation Report (1).

**Figure 3.**
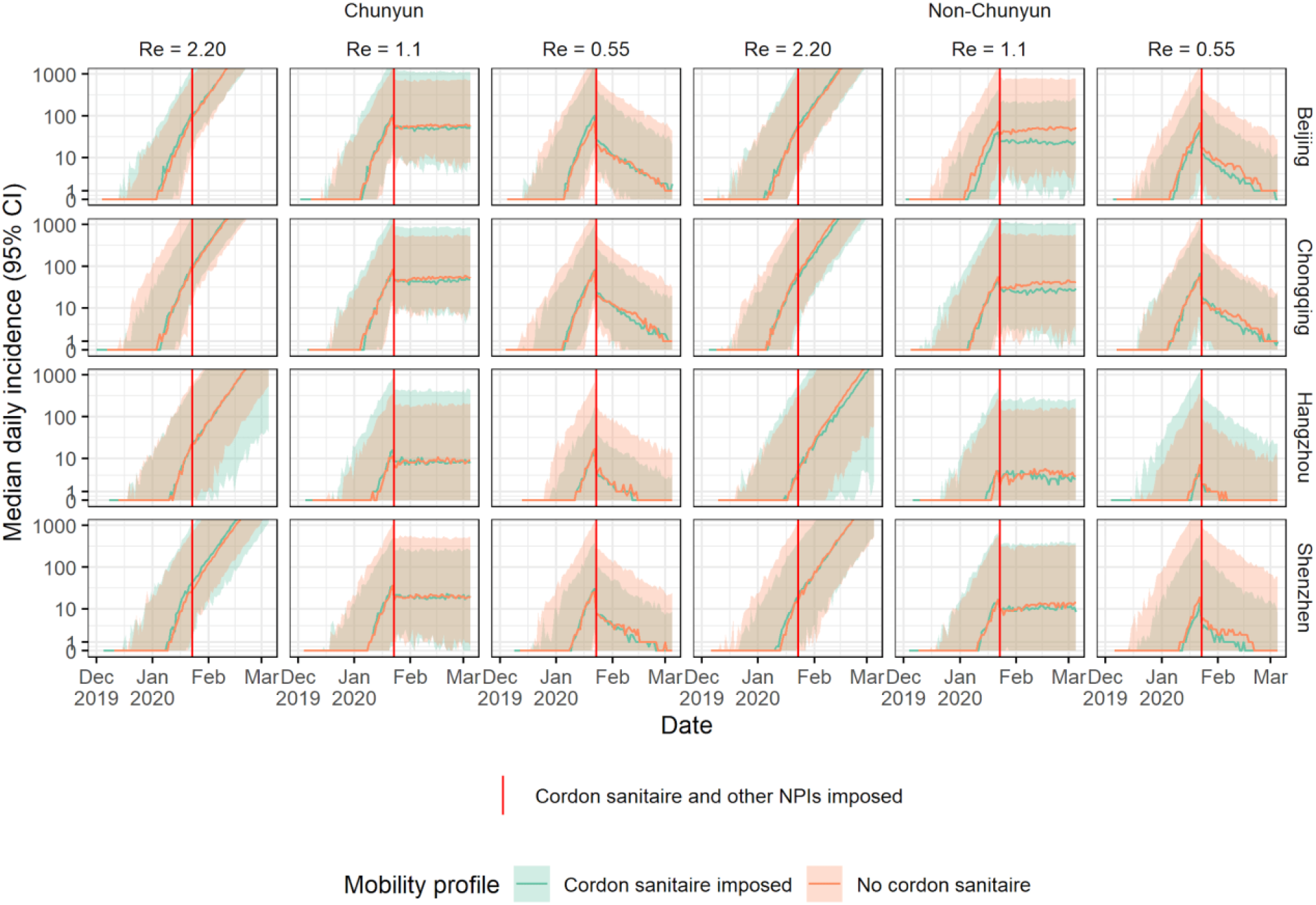
Median daily incidence of COVID-19 (log-scale, shaded areas indicate 95% confidence intervals) in the four cities of interest, for Chunyun vs. Non-Chunyun, cordon sanitaire imposed (orange) vs. no cordon sanitaire (green), and for varying values of the effective reproduction number R_e_, where R_e_ = 2.2 (no change, unmitigated local outbreak), reduced from 2.2 by 50% to 1.1 (mitigation of outbreak, R_e_>1), and 75% to 0.55 (suppression of outbreak, R_e_<1).

Similarly, no substantial differences were observed in the estimated cumulative number of infections by 1 March with and without *cordon sanitaire* in any of the four cities, after accounting for uncertainty resulting from the importation process and variability in the number of secondary cases resulting from an infected case (Figure 4).

**Figure 4.**
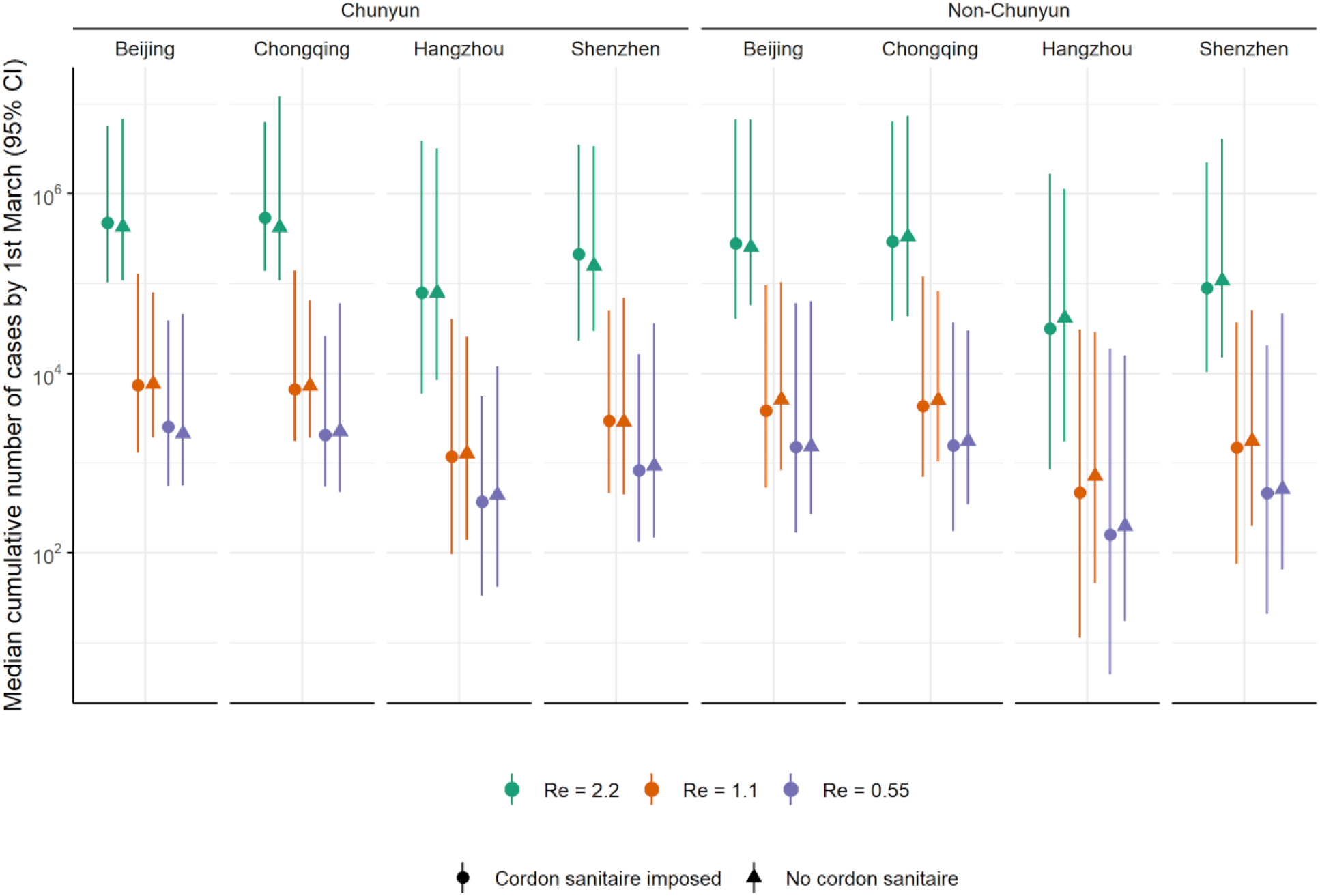
Median (and 95% CI) estimated cumulative number of infections on 1 March in the four cities of interest, Chunyun vs. Non-Chunyun, cordon sanitaire imposed vs. no cordon sanitaire, and for varying values of R_e_, where R = 2.2 (no change, unmitigated local outbreak), reduced from 2.2 by 50% to 1.1 (mitigation of outbreak, R>1), and 75% to 0.55 (suppression of outbreak, R<1).

## Discussion

By using publicly available mobility data to model the spread of the outbreak from Wuhan to other large population centres in China, we find that infected travellers from Wuhan likely led to local transmission in other major Chinese cities weeks before the *cordon sanitaire*. Cities with more travellers from Wuhan likely experienced higher incidence sooner. Modelling the trajectory of the outbreaks up to 1 March, in scenarios with and without the effect of the *cordon sanitaire*, we find no substantial differences in the cumulative number of infections generated. Furthermore, we found no evidence that the *Chunyun* travel period exacerbated the outbreak in these four cities.

By comparing *Chunyun* and non-*Chunyun* travel scenarios, no substantial difference was observed in terms of the cumulative number of infections generated by 1 Mar. This may be due to the consistent high volume of travel these cities receive from Wuhan year round, making it highly likely that infected travellers would still have arrived and seeded their own chains of transmission even during a period of regular travel volume. This however may differ in smaller cities which receive highly seasonal influxes of travellers from Wuhan relating to *Chunyun*.

The increase in mobility in 2020 compared to 2019 prior to LNY could be explained by a variety of factors, including year-to-year variations and potential factors related to COVID-19, such as the rumours of a rapidly growing outbreak and impending travel restrictions. In Northern Italy, a leaked COVID-19 plan might have driven thousands to flee south (20).

Many assumptions were made in the formulation of the travel scenarios. The Baidu Huiyan mobility values were presented on a relative index scale; we assumed a linear scaling factor of 50,000 travellers per unit. This was based on widely quoted estimates of people leaving Wuhan and the inter-city capacity of the travel network (7,10–13,21,22) (Supplementary Appendix 1). However, the index may represent a different number of travellers, or the scale may even be non-linear and the result of a more complex function, but without other evidence we assume linearity, as have other studies (7,12). If we chose a higher scaling factor, similar to ones used in other studies (11,12), it is likely that infected travellers would have arrived even earlier and in greater numbers to the four destination cities. Additionally, by reconstructing travel outflows for both dates outside of the observed range (22 Nov - 31 Dec) and simulated aspects of our scenarios, i.e. *Chunyun* affected travel days in non-*Chunyun* scenarios, the actual travel pattern may not have been accurately represented. Further assumptions were also made surrounding the pairwise travel flows, as observed data was only available for 2020, and the travel flows between Wuhan and each other prefecture-level city may have differed in 2019. We only considered Wuhan to be the sole source of infected individuals, and we only accounted for travellers making single-leg journeys to their destination. As such, we may underestimate the number of infected persons arriving by not considering the number of travellers which may have stopped in an intermediate location, become infected, and then arrived at the destination to seed local transmission, or indeed infected travellers arriving from outside of Wuhan. Hence most of our assumptions likely underestimated the number of travellers from Wuhan, and our conclusions would likely be the same even if the true number was higher. However, we also assumed that individuals would travel regardless of their infection status, which may overestimate the number of infections in destination cities.

As recent studies have shown (7,8,23), strict physical distancing measures soon decreased the effective reproduction number to 1 or less in Wuhan and other cities in China. By incorporating this decrease into our model, we find that *cordons sanitaires* alone, implemented after outbreaks were likely to be established in other cities, were likely ineffective in stopping outbreaks of COVID-19 in other major population centres. To have a greater impact, the *cordon sanitaire* would need to be implemented earlier, as investigated in Lai et al. 2020 (16), and be accompanied by other NPIs, such as general physical distancing and school and work closures (7,18). Similarly, it is unlikely that *cordons sanitaires* in other countries with well-established, geographically dispersed outbreaks will substantially delay COVID-19 spread. An open question is whether travel restrictions may be more efficacious to prevent or delay reintroductions after the lifting of other NPIs.

While earlier restrictions on travel from Wuhan may have had a larger impact, in countries with a high-degree of inter-city travel, it may be difficult to implement such highly disruptive travel restrictions at an early stage of the epidemic, before local transmission has already occurred in other cities. We find that local transmission in the four cities we studied (here defined as the probability of sustained transmission exceeding a 95% threshold) was most likely established between 2 January and 8 January; it was only on 8 January that the etiology of the “mystery pneumonia” (which was not yet confirmed to spread from person-to-person (25)) was determined as a novel coronavirus, and the first death occurred (26). It is difficult to see how the *cordon sanitaire* could have been justified any earlier, as almost every aspect of COVID-19 virology and epidemiology was unknown. Hence, it is likely that the sustained decline in COVID-19 incidence in other cities of China several months into the outbreak is primarily due to other public health measures to reduce the disease transmissibility, i.e, to reduce the reproduction number to 1 or below (8,18,23). The *cordon sanitaire* may have been more efficacious in delaying outbreaks internationally, as the relative number of travellers is orders of magnitude lower (7,27); the same may also apply to lower-traffic destinations from Wuhan within China, such as small cities geographically distant from Wuhan, as observed in Tian et al. 2020 (7). We found a mixed picture in these cities, where the *cordon sanitaire* may have been more efficacious at delaying or preventing outbreaks (Figure S1). However, COVID-19 transmission dynamics may differ in comparison to big cities, and as such we chose to focus on the effect of travel restrictions in large cities with large volumes of travel from Wuhan, where data on *R* and *k* from the early outbreak in Wuhan are likely generalisable. Furthermore, these destinations with low traffic from Wuhan are more likely to be seeded by outbreaks in other, comparatively closer, large cities first. Hence, our assumption of a single outbreak source would have been much less realistic.

Our estimated dates of introduction in other cities are earlier than those observed (1) and reported in other studies (28). This is due in part to correction for underreporting, both by using the estimated daily prevalence in Wuhan from Kucharski et al. 2020 (8), which is significantly higher than the confirmed number of cases (20), and by not relying on reported cases in other provinces. The effect of underreporting is likely more pronounced early in the outbreak prior to a well-defined case definition or widespread testing (14). Hence, reconstructing the early outbreak through a simulation approach was more appropriate in this setting.

We concur with Tian et al. 2020 (7) that prohibiting travel alone did not act to reduce the number of COVID-19 infections in four major cities outside of Wuhan or Hubei, and that other local control measures were likely instrumental in reducing incidence. Likewise, Kraemer et al. 2020 (28) conclude that while a decrease in the growth rate was observed in large cities after the *cordon sanitaire* was imposed, this is difficult to disentangle from local control measures.

In conclusion, the introduction of *cordon sanitaire*-type travel restrictions around a COVID-19 epidemic centre after community transmission is already occurring in other well-connected population centres on its own likely has little effect on altering their epidemic trajectories. Stringent NPIs in cities are more likely to have a bigger impact in reducing incidence and pressure on healthcare systems. Further research should examine the role of travel restrictions during the partial lifting of NPIs across China and elsewhere.

## Data Availability

The mobility data was sourced from Baidu Haiyan migration dashboard. The code for this analysis is available on GitHub.

https://qianxi.baidu.com/

https://github.com/bquilty25/wuhan_travel_restrictions

## Data availability

The mobility data was sourced from Baidu Haiyan migration dashboard at https://qianxi.baidu.com/

The code for this analysis is available on GitHub at https://github.com/bquilty25/wuhan_travel_restrictions

## Acknowledgements

BJQ, CD, YL, KP, PK, RME and MJ conceived the study. BJQ and CD designed and programmed the model and made the figures. YL, HG, TWR and CIJ provided the data. YL, HG, TWR, CIJ, KP, CABP, SC, SF, PK, REM and MJ consulted on the analyses. BJQ, CD, YL and MJ wrote the manuscript. The CMMID COVID-19 working group members contributed to processing, cleaning, and interpretation of data, interpreted the study findings, contributed to the manuscript, and approved the work for publication. All authors interpreted the findings, contributed to writing the manuscript, and approved the final version for publication.

BJQ, CD, YL, and MJ were funded by the National Institute for Health Research (NIHR; 16/137/109). YL, PK, KP and MJ were funded by the Bill & Melinda Gates Foundation (INV-003174). HG was funded by the Department of Health and Social Care (ITCRZ 03010). TWR was funded by the Wellcome Trust (206250/Z/17/Z). CIJ was funded by the Global Challenges Research Fund (ES/P010873/1). CABP was funded by the NTD Modelling Consortium by the Bill and Melinda Gates Foundation (OPP1184344). SF and SC were funded by the Sir Henry Dale Fellowship (208812/Z/17/Z). RME was funded by Health Data Research UK (MR/S003975/1). This research was partly funded by the NIHR (16/137/109) using aid from the UK Government to support global health research. The views expressed in this publication are those of the author(s) and not necessarily those of the NIHR or the UK Department of Health and Social Care.

We would like to acknowledge the other members of the London School of Hygiene & Tropical Medicine CMMID COVID-19 modelling group, who contributed to this work. Their funding sources are as follows: Jon C Emery and Rein M G J Houben (European Research Council Starting Grant, Action Number #757699); Megan Auzenbergs and Kathleen O’Reilly (Bill and Melinda Gates Foundation, OPP1191821); Nicholas Davies (NIHR HPRU-2012-10096); Emily S Nightingale (Bill and Melinda Gates Foundation, OPP1183986); Kevin van Zandvoort (Elrha’s Research for Health in Humanitarian Crises [R2HC] Programme, UK Government [Department for International Development], Wellcome Trust, and NIHR); Thibaut Jombart (Research Public Health Rapid Support Team, NIHR Health Protection Research Unit Modelling Methodology); Arminder K Deol; W John Edmunds; Joel Hellewell, Sam Abbott, James D Munday, Nikos I Bosse and Sebastian Funk (Wellcome Trust 210758/Z/18/Z); Fiona Sun (NIHR; 16/137/109); Akira Endo (The Nakajima Foundation; The Alan Turing Institute); Alicia Rosello (NIHR: PR-OD-1017-20002); Amy Gimma (Global Challenges Research Fund ES/P010873/1); Simon R Procter (Bill and Melinda Gates Foundation, OPP1180644); Graham Medley (NTD Modelling Consortium by the Bill and Melinda Gates Foundation (OPP1184344); Adam J Kucharski (Wellcome Trust, 206250/Z/17/Z), and Gwen Knight (UK Medical Research Council, MR/P014658/1).

## Competing interests

Akira Endo received a research grant from Taisho Pharmaceutical Co., Ltd. We declare no other competing interests.

Neither patients nor the public were involved with the design, conduct, reporting, or dissemination plans of our research. As this work is a simulation study, there are no participants to which we can disseminate the results of this research.

## Supplementary Appendix

### 1. Estimating the scaling factor

To estimate the absolute number of daily travellers leaving Wuhan from Baidu’s migration index, we needed a suitable scaling factor to convert the index score to the absolute number of travellers. In lieu of other evidence, we assumed this relationship to be linear cohering with other studies (7,12). We synthesised estimates from a number of sources (Table S1) in order to select the most viable result. In each case the scaling factor was calculated using the following equation:

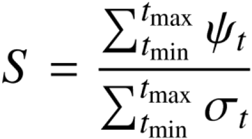

Where the sum of the daily estimated number of travellers *ψ*_*t*,_leaving Wuhan for the dates *t*_*min*_ to *t*_*max*_, divided by the sum of the daily outflow index from Wuhan σ_t_, for the same date range, equals the scaling factor *S*.

**Table S1.** Various scaling factors calculated from different sources.

| Reference | Date range ($t_{\min}$ to $t_{\max}$) | Sum of traveller numbers leaving Wuhan ($\psi_t$) | Sum of Baidu travel index leaving Wuhan ($\sigma_t$) | Estimated scaling factor (S) |
| --- | --- | --- | --- | --- |
| Tian (2020) (7) | Jan 11 - Jan 25 | 4,325,563* | 105.69 | 40,926.89 |
| Sanche (2020) (10) | Jan 10 - Jan 23 | 5,000,000 | 107.12 | 46,676.62 |
| News report (2020) (13) | Jan 10 - Jan 20 | 4,098,600 | 73.40 | 55,839.24 |
| Cao (2020) (11) | Jan 16 - Jan 22 | 7,014,199* | 58.45 | 120,003.40 |
| Zhou (2020) (12) | Unknown | Unknown | Unknown | 138,412.00 |

\* Based on data extracted from figures, subject to slight error.

Combining evidence from the first three sources in Table S1, we chose a scaling factor of 50,000. This assumes each unit of Baidu’s migration index corresponds to 50,000 outbound travellers. This produced the most reasonable outbound travel volume estimates, using scaling factors of 120,003.40 and 138,412, found in Cao (2020) and Zhou (2020) respectively, yielded unrealistically large travel magnitudes. These scaling factors would suggest that on Beijing’s single busiest day of *Chunyun* (23 Jan, 2020), in excess of 2.8 million people left the city. This is substantially larger than Beijing’s maximum daily outbound travel capacity by air and rail, which is estimated to be 920,000 daily passengers (21,22). This estimate does not consider passengers traveling by road, however this form of transportation accounts for a relatively small proportion of the total inter-prefecture travel.

### 2. Estimating number of infected travellers

The number of travellers arriving in each city from Wuhan is summarised as:

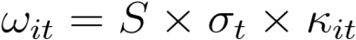

Where *S* is the scaling factor, *σ*_*t*_ is the total daily outflow index from Wuhan, *κ*_*it*_ is the daily proportion of outflow entering each city *i*, and *ω*_*it*_ is the daily number of total arrivals from Wuhan in city *i*.

The number of daily infected arrivals to a given prefecture *i* is simulated by making 200 draws from a Poisson process:

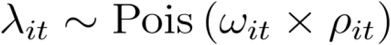

Where *ω*_*it*_ is the daily estimated travel from Wuhan to prefecture *i* on day *t, ρ*_*it*_ is the daily prevalence in Wuhan, and *λ*_*it*_ is the number of infected individuals arriving per day.

### 3. Travel flow scenario formulation

The observed travel outflow from Wuhan in 2019 and 2020 were matched by the date of the Lunar New Year in 2020 so as to align the *Chunyun* travel patterns. Each scenario is driven by differences in the parameters used to estimate the total daily number of travellers arriving from Wuhan in a given prefecture-level city. In all scenarios the scaling factor was assumed to be constant at 50,000. Differences between scenarios are summarised in the table and equations below:

**Table S2.** Parameters used to estimate the total number of travellers leaving Wuhan and entering other prefecture-level cities for each scenario.

| Scenario and description | Daily outflow from Wuhan ($\sigma_t$) | Daily proportion of travellers leaving Wuhan and entering each prefecture-level city ($\kappa_{it}$) |
| --- | --- | --- |
| **Scenario 1** - Chunyun & cordon sanitaire | 22 Nov - 31 Dec: $\overline{\sigma}_t^*$<br>1 Jan - 1 Mar: $\sigma_t$ (Observed 2020) | 22 Nov - 31 Dec: $\overline{\kappa}_{it}^\dagger$<br>1 Jan - 1 Mar: $\kappa_{it}$ (Observed 2020) |
| **Scenario 2** - Chunyun & no cordon sanitaire | 22 Nov - 31 Dec: $\overline{\sigma}_t^*$<br>1 Jan - 1 Mar: $\sigma_t$ (Observed 2019<sup>^</sup>) | 22 Nov - 31 Dec: $\overline{\kappa}_{it}^\dagger$<br>1 Jan - 19 Jan: $\kappa_{it}$ (Observed 2020)<br>20 Jan - 1 Mar: $\overline{\kappa}_{it}^\dagger$ |
| **Scenario 3** - No Chunyun & cordon sanitaire | 22 Nov - 5 Jan: $\overline{\sigma}_t^*$<br>6 Jan - 10 Jan: $\sigma_t$ (Observed 2019<sup>^</sup>)<br>11 Jan - 23 Jan: $\overline{\sigma}_t^*$<br>24 Jan - 1 Mar: $\sigma_t$ (Observed 2020) | 22 Nov - 23 Jan: $\overline{\kappa}_{it}^\dagger$<br>24 Jan - 1 Mar: $\kappa_{it}$ (Observed 2020) |
| **Scenario 4** - No Chunyun & no cordon sanitaire | 22 Nov - 5 Jan: $\overline{\sigma}_t^*$<br>6 Jan - 10 Jan: $\sigma_t$ (Observed 2019<sup>^</sup>)<br>11 Jan - 7 Feb: $\overline{\sigma}_t^*$<br>Feb 8 - 1 Mar: $\sigma_t$ (Observed 2019<sup>^</sup>) | 22 Nov - 1 Mar: $\overline{\kappa}_{it}^\dagger$ |

<sup>^</sup> Equivalent Chunyun dates aligned to the 2020 calendar.
\* See equation 1.
<sup>†</sup> See equation 2.

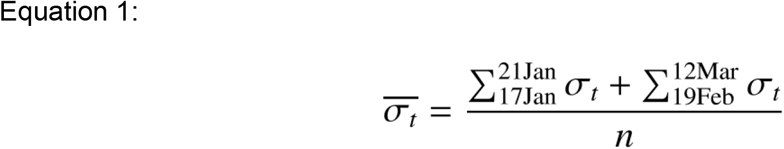

The above equation estimates the mean daily outflow index from Wuhan in 2019 for the following dates; 17 Jan - 21 Jan and 19 Feb - 12 Mar. These dates are understood to be days of regular travel volume, and as such can be used to construct an estimate of an average travel flow for a representative non-*Chunyun* period.

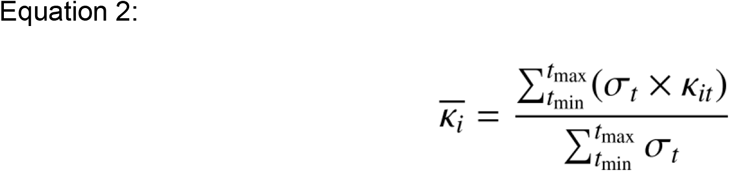

Equation 2 approximates the general daily proportion of travellers leaving Wuhan and entering a given city *i*. For the length of the study period (*t*_*min*_ to *t*_*max*_), we take the sum of the estimated travel flow leaving Wuhan and entering city *i* (*σ*_*t*_ *× κ*_*it*_) and divide it by the sum of the total outflow from Wuhan *σ*_*t*_ over the same period. This was a key assumption as the pairwise travel flows between Wuhan and each other prefecture-level city was only available between 1 Jan - 1 Mar, 2020. Therefore this approximation of general flow magnitude was used for both out of date ranges (22 Nov - 31 Dec) and simulated aspects of our scenarios i.e. *Chunyun* affected travel days in non-*Chunyun* scenarios.

### 4. Probability of sustained transmission (outbreak threshold)

The probability of sustained transmission was calculated using methods detailed in the Supplement of Hartfield and Alizon 2013 (30), which we will briefly summarise here.

Given a secondary case distribution with mean *R* and dispersion parameter *k*, the individual probability of a outbreak *q* can be numerically solved by:

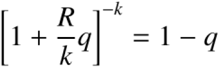

The number of individuals required such that the probability at least one of them causes an outbreak *i* is then:

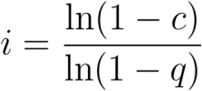

Where *c* (the outbreak threshold) was chosen as 0.95.

The impact of travel restrictions was assessed by comparing the daily probability of an outbreak occurring *o* for 2020 (restrictions imposed) and 2019 (“business-as-usual”).

### 5. Supplementary figures and tables

**Figure S1.**
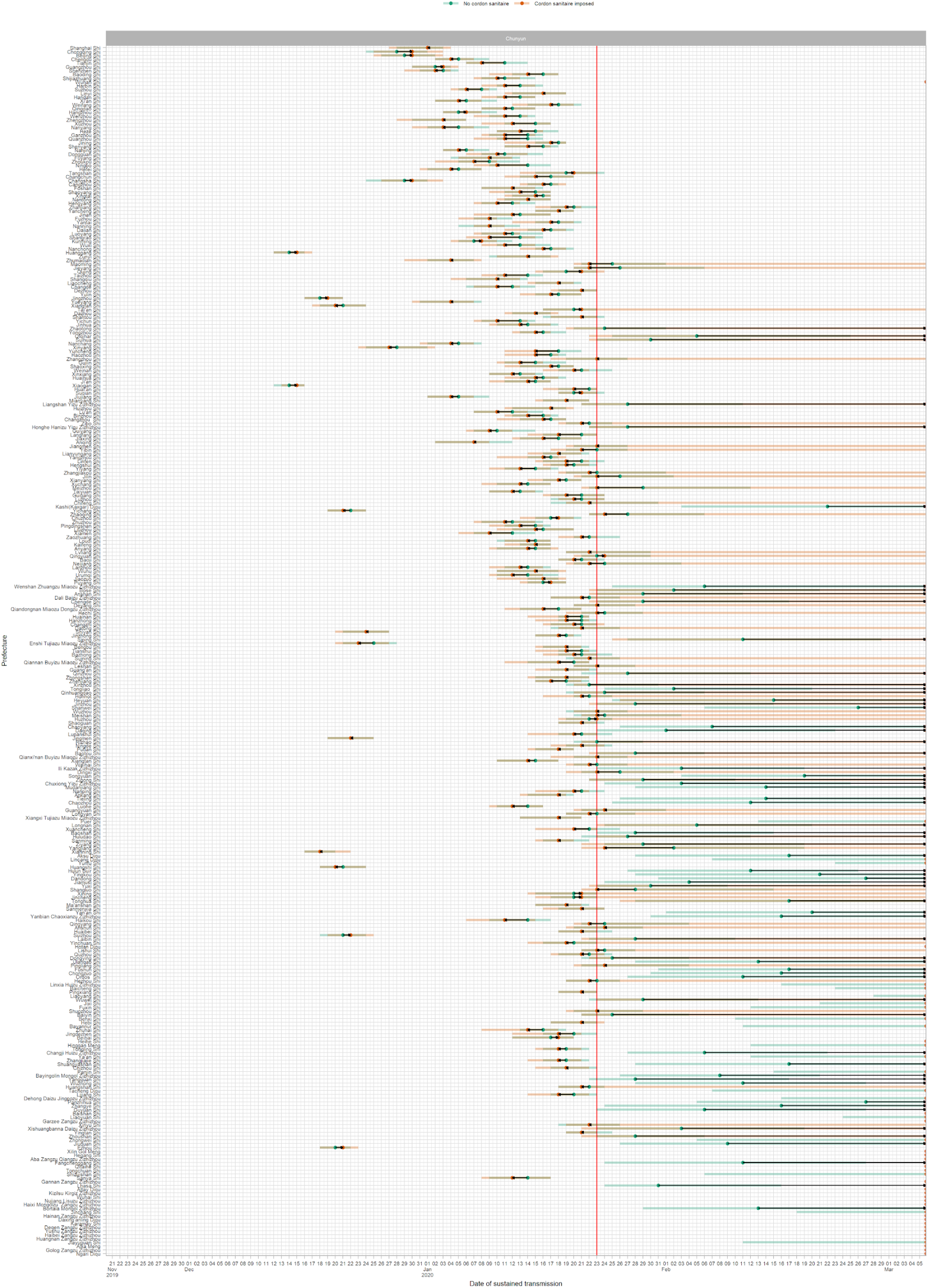
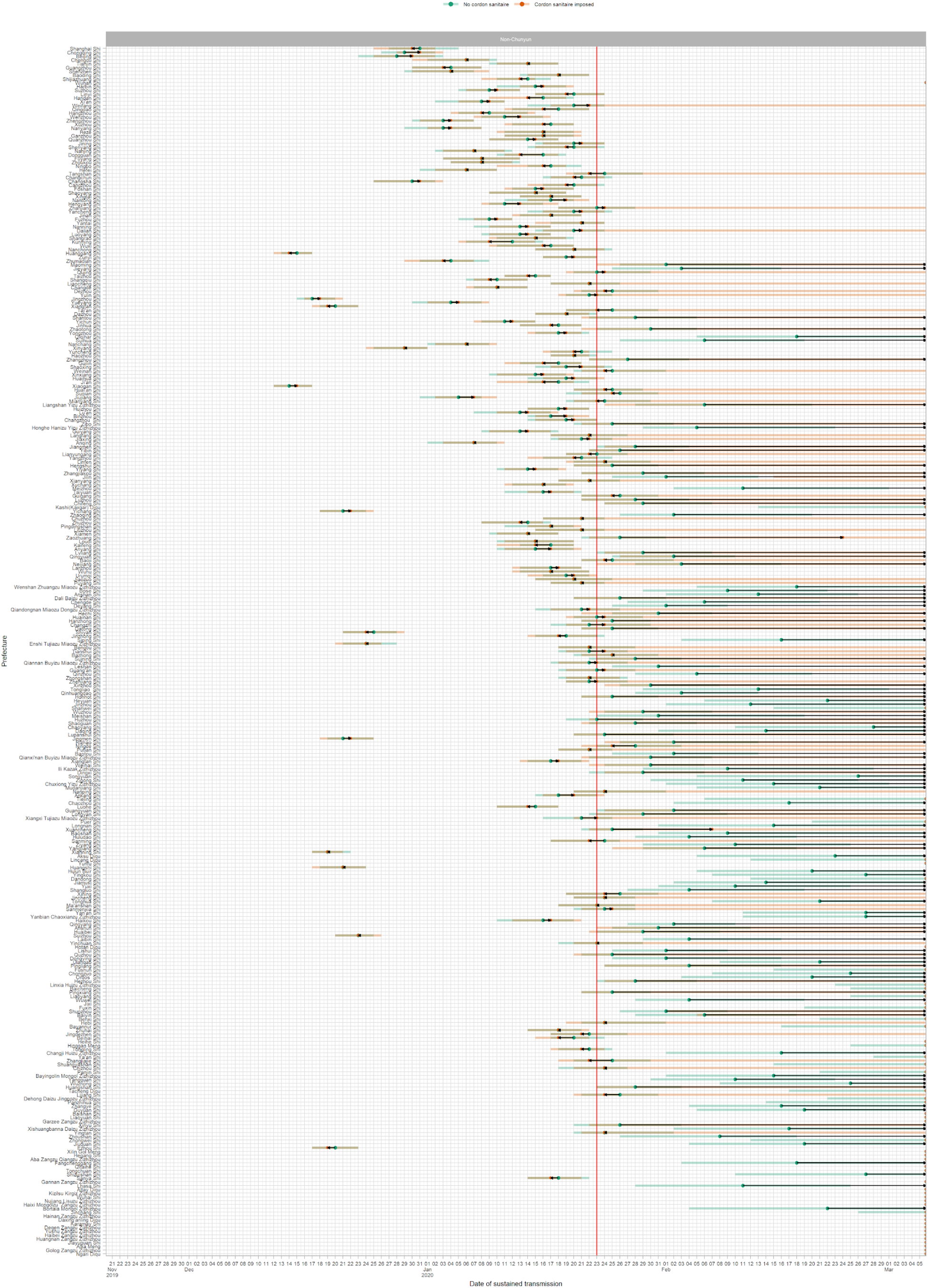
Date at which the mean probability of sustained transmission breaches 95% (30) for cordon sanitaire imposed (orange) vs no cordon sanitaire (green) for Chunyun (top panel) and Non-Chunyun (lower panel) travel patterns for each prefecture given travel patterns from Wuhan. Red vertical line indicates the date the cordon sanitaire imposed. Black lines with arrows indicate time difference between scenarios; arrows pointing right indicate delay, arrows pointing left indicate advance. Points on the right limit of the graph indicate that no outbreak has occurred. Prefectures sorted by population. Outbreak probability calculated with R0=2.2 and k=0.54.

**Figure S2.**
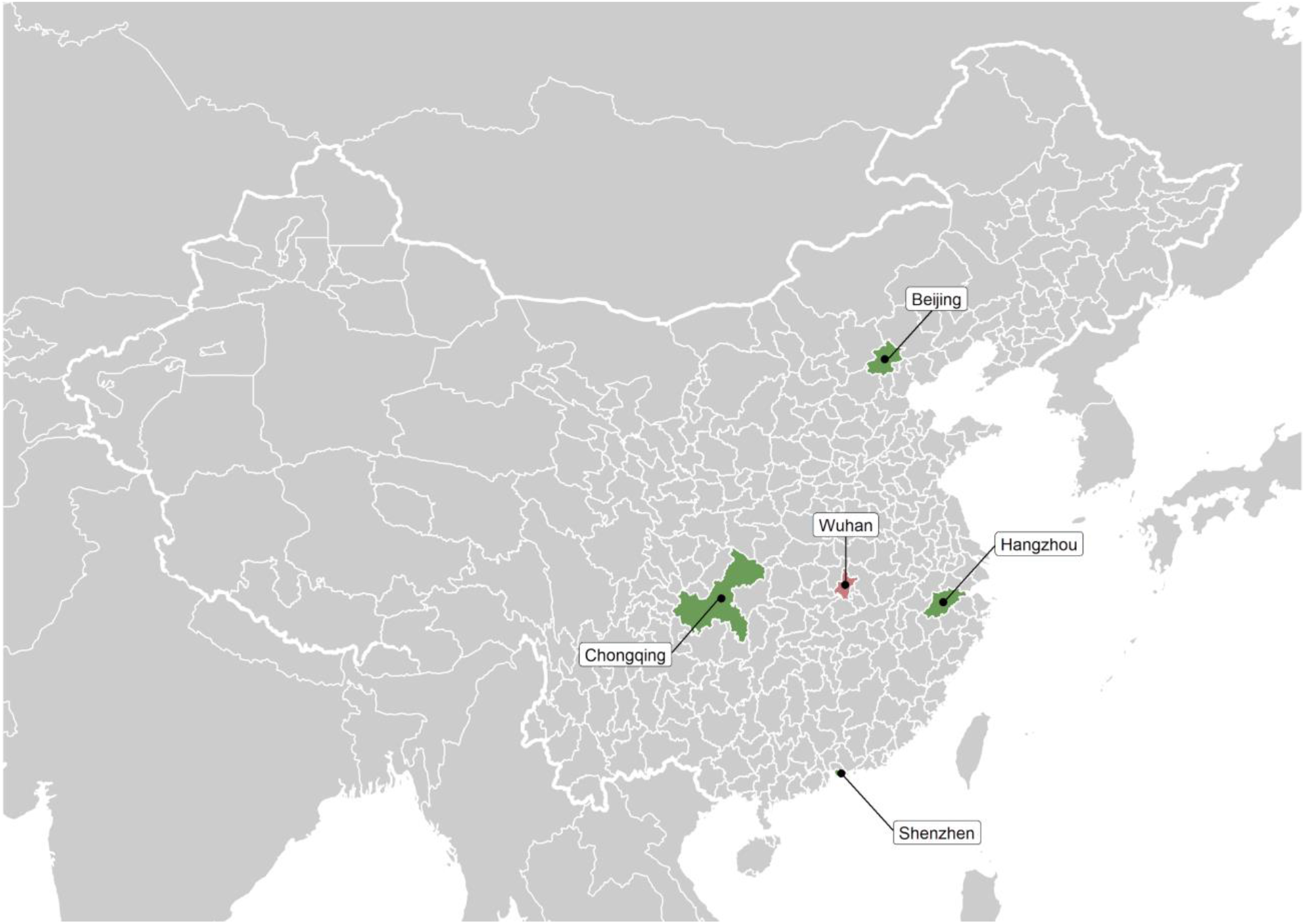
Location of Wuhan (centre, pink) and the four cities of interest (green) in mainland China.

**Table S3.** Sensitivity analysis for overdispersion parameter k. k = 0.16 (SARS-like), k = 2 (H1N1-like). Dates at which the probability of sustained transmission breached the 95% threshold for the four prefecture-level cities of interest in each of the 4 scenarios.

| City | $k$ | Date of probability of sustained transmission exceeding 95% threshold (median, 95% CI) | | | |
| --- | --- | --- | --- | --- | --- |
|  |  | Chunyun |  | Non-Chunyun |  |
|  |  | Cordon sanitaire imposed<br>(Scenario 1) | No cordon sanitaire<br>(Scenario 2) | Cordon sanitaire imposed<br>(Scenario 3) | No cordon sanitaire<br>(Scenario 4) |
| Beijing | 0.16 | 06 Jan (01 Jan - 10 Jan) | 06 Jan (31 Dec - 11 Jan) | 09 Jan (29 Dec - 15 Jan) | 09 Jan (30 Dec - 16 Jan) |
| Beijing | 0.54 | 02 Jan (22 Dec - 07 Jan) | 02 Jan (22 Dec - 10 Jan) | 02 Jan (22 Dec - 11 Jan) | 03 Jan (23 Dec - 12 Jan) |
| Beijing | 2 | 30 Dec (17 Dec - 05 Jan) | 31 Dec (18 Dec - 06 Jan) | 30 Dec (18 Dec - 09 Jan) | 31 Dec (17 Dec - 10 Jan) |
| Chongqing | 0.16 | 07 Jan (02 Jan - 10 Jan) | 08 Jan (02 Jan - 12 Jan) | 09 Jan (02 Jan - 15 Jan) | 08 Jan (31 Dec - 14 Jan) |
| Chongqing | 0.54 | 02 Jan (22 Dec - 08 Jan) | 02 Jan (22 Dec - 09 Jan) | 02 Jan (24 Dec - 10 Jan) | 02 Jan (22 Dec - 11 Jan) |
| Chongqing | 2 | 29 Dec (19 Dec - 07 Jan) | 30 Dec (15 Dec - 07 Jan) | 29 Dec (17 Dec - 09 Jan) | 29 Dec (15 Dec - 07 Jan) |
| Hangzhou | 0.16 | 13 Jan (06 Jan - 20 Jan) | 16 Jan (09 Jan - 20 Jan) | 20 Jan (11 Jan - Inf) | 19 Jan (12 Jan - 26 Jan) |
| Hangzhou | 0.54 | 08 Jan (31 Dec - 16 Jan) | 10 Jan (01 Jan - 17 Jan) | 12 Jan (31 Dec - 21 Jan) | 14 Jan (31 Dec - 23 Jan) |
| Hangzhou | 2 | 06 Jan (24 Dec - 14 Jan) | 06 Jan (22 Dec - 15 Jan) | 09 Jan (25 Dec - 18 Jan) | 08 Jan (26 Dec - 20 Jan) |
| Shenzhen | 0.16 | 10 Jan (05 Jan - 17 Jan) | 11 Jan (04 Jan - 17 Jan) | 15 Jan (06 Jan - 20 Jan) | 14 Jan (05 Jan - 20 Jan) |
| Shenzhen | 0.54 | 05 Jan (29 Dec - 13 Jan) | 06 Jan (26 Dec - 14 Jan) | 08 Jan (29 Dec - 16 Jan) | 08 Jan (27 Dec - 16 Jan) |
| Shenzhen | 2 | 03 Jan (18 Dec - 10 Jan) | 02 Jan (18 Dec - 11 Jan) | 05 Jan (19 Dec - 15 Jan) | 04 Jan (16 Dec - 14 Jan) |

